# Risk of myocarditis following sequential COVID-19 vaccinations by age and sex

**DOI:** 10.1101/2021.12.23.21268276

**Authors:** Martina Patone, Xue W Mei, Lahiru Handunnetthi, Sharon Dixon, Francesco Zaccardi, Manu Shankar-Hari, Peter Watkinson, Kamlesh Khunti, Anthony Harnden, Carol AC Coupland, Keith M. Channon, Nicholas L Mills, Aziz Sheikh, Julia Hippisley-Cox

**Affiliations:** Nuffield Department of Primary Health Care Sciences, University of Oxford; Wellcome Centre for Human Genetics, University of Oxford; Leicester Real World Evidence Unit, Diabetes Research Centre, University of Leicester, Leicester, UK; Usher Institute, University of Edinburgh, Edinburgh, UK; Centre for Inflammation Research, University of Edinburgh; School of Immunology and Microbial Sciences, King’s College London; Nuffield Department of Clinical Neurosciences, University of Oxford; NIHR Biomedical Research Centre, Oxford University Hospitals NHS Trust; School of Medicine, University of Nottingham; British Heart Foundation Centre of Research Excellence, NIHR Oxford Biomedical Research Centre, Radcliffe Department of Medicine, University of Oxford, John Radcliffe Hospital, Oxford, UK; BHF/University Centre for Cardiovascular Science, University of Edinburgh, Edinburgh, UK

**Author notes:** **Correspondence to:** Professor Julia Hippisley-Cox, Nuffield Department of Primary Care Health Sciences, Radcliffe Observatory Quarter, Woodstock Road.

## Abstract

In an updated self-controlled case series analysis of 42,200,614 people aged 13 years or more, we evaluate the association between COVID-19 vaccination and myocarditis, stratified by age and sex, including 10,978,507 people receiving a third vaccine dose. Myocarditis risk was increased during 1-28 days following a third dose of BNT162b2 (IRR 2.02, 95%CI 1.40, 2.91). Associations were strongest in males younger than 40 years for all vaccine types with an additional 3 (95%CI 1, 5) and 12 (95% CI 1,17) events per million estimated in the 1-28 days following a first dose of BNT162b2 and mRNA-1273, respectively; 14 (95%CI 8, 17), 12 (95%CI 1, 7) and 101 (95%CI 95, 104) additional events following a second dose of ChAdOx1, BNT162b2 and mRNA-1273, respectively; and 13 (95%CI 7, 15) additional events following a third dose of BNT162b2, compared with 7 (95%CI 2, 11) additional events following COVID-19 infection. An association between COVID-19 infection and myocarditis was observed in all ages for both sexes but was substantially higher in those older than 40 years. These findings have important implications for public health and vaccination policy.

**Funding:** Health Data Research UK.

## MAIN

Our recent article on the association between COVID-19 vaccination and myocarditis generated considerable scientific, policy and public interest [1]. It added to evidence emerging from multiple countries that have linked exposure to BNT162b2 messenger RNA vaccine with acute myocarditis [2-8]. In the largest and most comprehensive analysis to date, we confirmed prior findings and reported an increase in hospital admission or death from myocarditis following three different types of vaccine including both mRNA and adenoviral vaccines.

Importantly, we also demonstrated that across the entire vaccinated population in England, the risk of myocarditis following vaccination was small compared to the risk following a positive severe acute respiratory syndrome coronavirus 2 (SARS-CoV-2) test [1]. However, myocarditis is more common in younger persons and in males in particular [9, 10]. Additional analyses stratified by both age and sex and following a third vaccine dose were requested as vaccine campaigns are rapidly being extended to include children and young adults. Furthermore, given the consistent observation that the risk of myocarditis is higher following the second dose of vaccine compared to the first dose [1, 11], there is an urgent need to evaluate the risk associated with a third dose as booster programmes are accelerated internationally to combat the omicron variant [12].

We therefore extended our analysis to include persons aged 13 years or more and those receiving a third dose to further evaluate the association between COVID-19 vaccination or infection and myocarditis, stratified by age and sex.

In brief, we used the NHS Immunisation Management Service (NIMS) database, which includes data for all people receiving a COVID-19 vaccine in England. We linked individual patient data to national data for hospital admission, mortality and SARS-CoV-2 testing to examine associations between exposures to the first, second or third dose of ChAdOx1, BNT162b2 or mRNA-1273 vaccine, or a positive SARS-CoV-2 test before or after vaccination, and hospital admission or death from myocarditis. The self-controlled case series (SCCS) method [13, 14] compares the incidence rate of myocarditis in exposed and unexposed periods within individuals implicitly controlling for within person covariates. The incidence rate ratio (IRR) is calculated for hospital admission or death in a 1-28 day risk period after vaccination or a positive test, compared to baseline periods. The IRR was calculated following stratification by sex and age in those younger or older than 40 years.

Between December 1, 2020, to November 15, 2021 a total of 42,200,614 people were vaccinated with at least one dose of ChAdOx1 (n=20,646,456), BNT162b2 (n=20,391,600) or mRNA-1273 (n=1,162,558) in England (Supplementary Table 1). Of these, 38,347,981 received two doses of either ChAdOx1 (n=20,059,058), BNT162b2 (n=17,294,004) or mRNA-1273 (n=1,039,919) and 10,978,507 people received a third dose of ChAdOx1 (n=35,608), BNT162b2 (n=10,599,183) or mRNA-1273 (n=343,716). Amongst people receiving at least one vaccine dose, 5,185,772 (12.3%) tested positive for SARS-CoV-2; 2,834,579 (54.7%) prior to vaccination, 698,993 (13.5%) after a first vaccine dose, 1,604,087 (30.9%) after a second vaccine dose and 48,113 (0.9%) after a third vaccine dose. Of the 42,200,614 persons included in the study population, 2,539 (0.006%) were hospitalised or died from myocarditis during the study period; 552 (0.001%) of these events occurred during 1-28 days following any dose of vaccine (Supplementary Table 2).

Over the 1-28 days post vaccination, we observed an association with the first dose of ChAdOx1 (IRR 1.27, 95%CI 1.05, 1.55) and BNT162b2 (IRR 1.37, 95%CI 1.12, 1.67), but not mRNA-1273 (IRR 1.80, 95%CI 0.91, 3.58; Table 1 and Extended Figure 1). Following a second dose, the risk was higher with mRNA-1273 (IRR 13.71, 95%CI 8.46, 22.20) compared to BNT162b2 (IRR 1.60, 95%CI 1.31, 1.97). No association with a second dose of ChAdOx1 was found. An association after a third dose was only observed for BNT162b2 (IRR 2.02, 95%CI 1.40, 2.91). No myocarditis events occurred 1-28 days after a third dose in the small number of persons receiving ChAdOx1 or mRNA-1273 vaccine. The risk of myocarditis was increased in the 1-28 days following a SARS-CoV-2 positive test (IRR 8.40, 95%CI 6.89, 10.25).

**Table 1:**
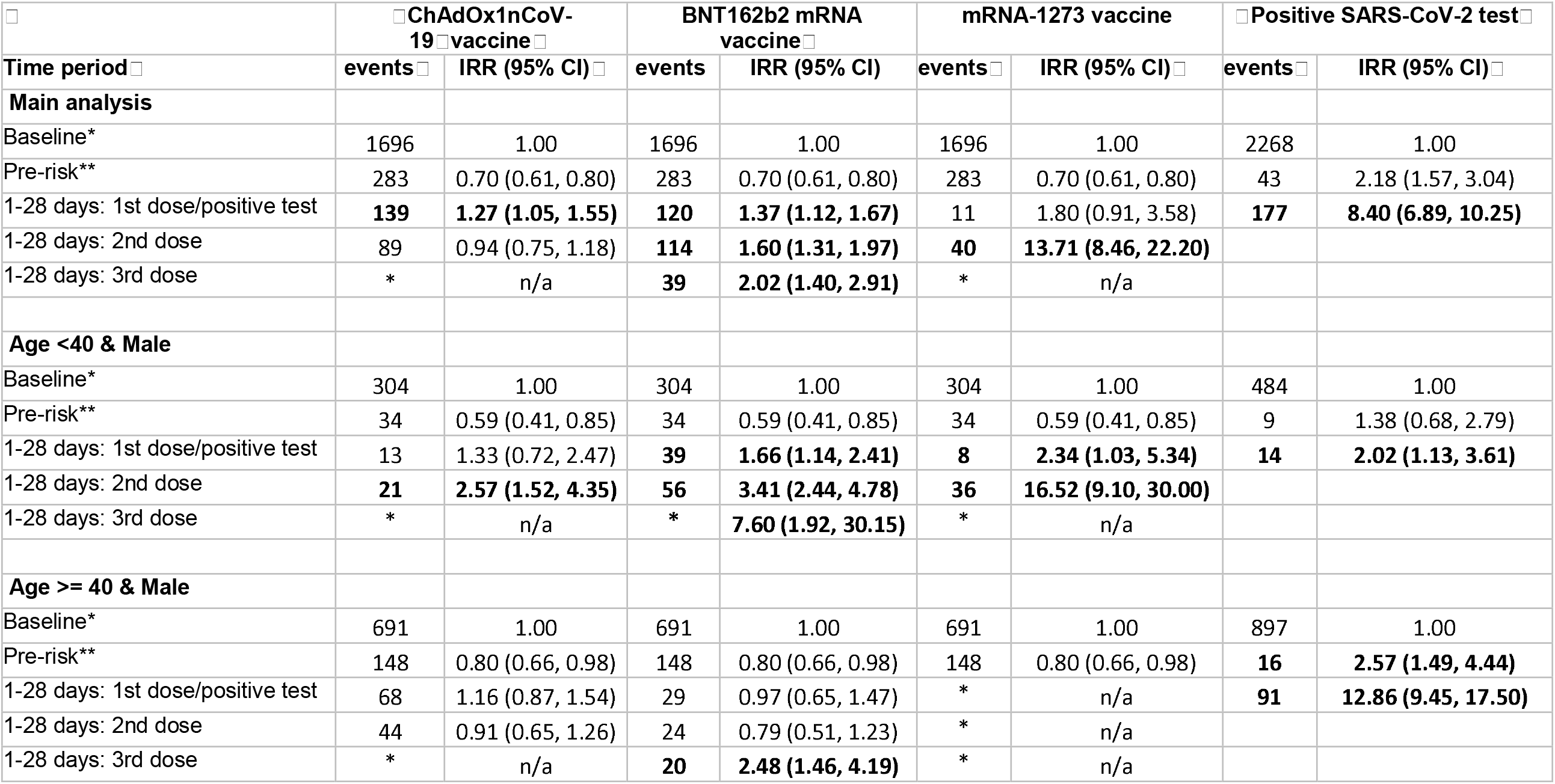
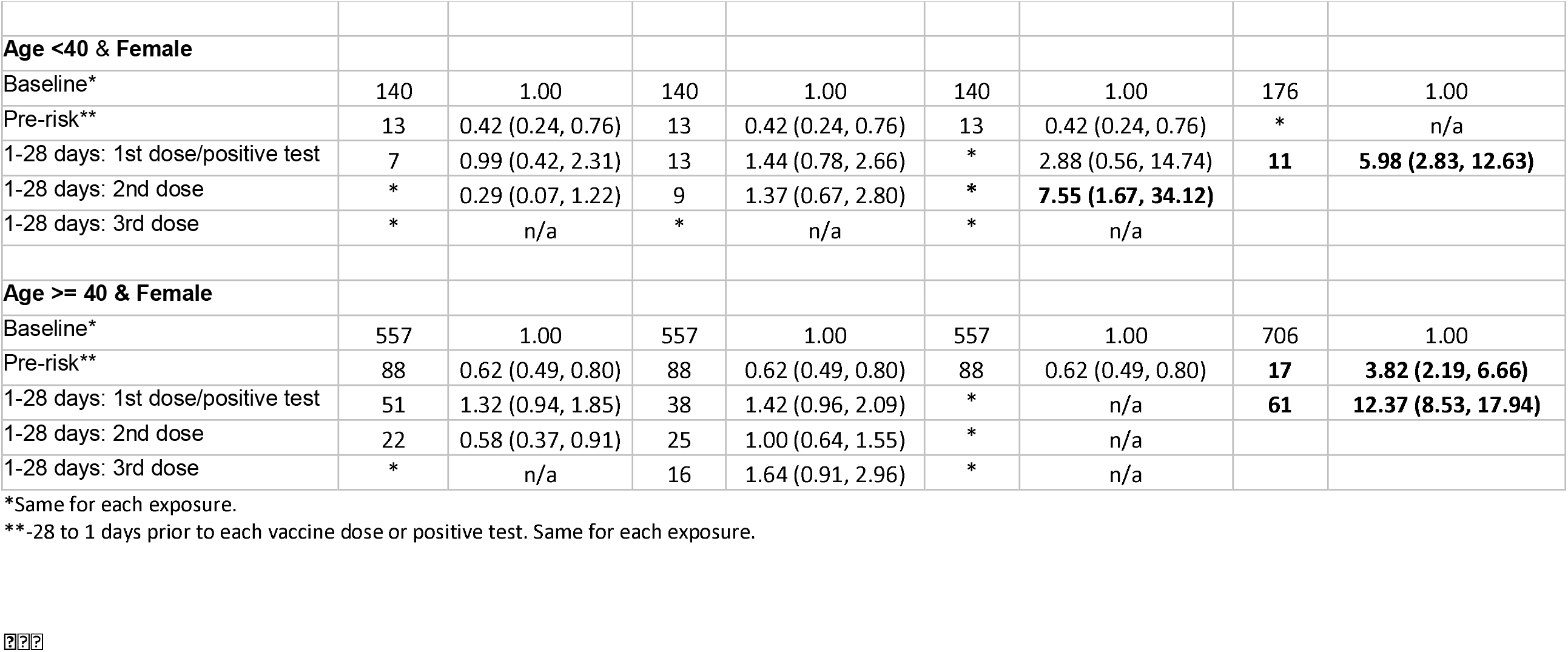
**Incidence rate ratios (IRR 95% CI) for main analysis and by age group (aged 40 or younger, older than 40) and sex (female and male) for the outcomes in pre-defined risk periods immediately before and after exposure to vaccination and before and after a positive SARS-CoV-2 test result, adjusted for calendar time from December 1 2020 to November 15 2021 (cells with * are suppressed as counts < 5). Day 0 of each exposure has been removed due to small numbers**.

In males aged less than 40 years, we observed an increased risk of myocarditis in the 1-28 days following a first dose of BNT162b2 (IRR 1.66, 95%CI 1.14, 2.41) and mRNA-1273 (IRR 2.34, 95%CI 1.03, 5.34); after a second dose of ChAdOx1 (2.57, 95%CI 1.52, 4.35), BNT162b2 (IRR 3.41, 95% CI 2.44, 4.78) and mRNA-1273 (IRR 16.52, 95%CI 9.10, 30.00); after a third dose of BNT162b2 (IRR 7.60, 95%CI 2.44, 4.78); and following a SARS-CoV-2 positive test (IRR 2.02, 95%CI 1.13, 3.61; Extended Figure 1 and Table 1). In older males, the risk of myocarditis was increased 1-28 days following a third dose of BNT162b2 vaccine (IRR 2.48, 95%CI 1.46, 4.19) and following a positive test (IRR 5.98, 95%CI 2.83, 12.63).

In females aged less than 40 years, we only observed an increased risk of myocarditis in the 1-28 days following a second dose of mRNA-1273 vaccine (IRR 7.55, 95%CI 1.67, 34.12; Figure 1). However, the numbers of events were small. In older females, we found no association between myocarditis and vaccination. Supplementary Table 4 shows IRRs per week following exposure.

**Figure 1:**
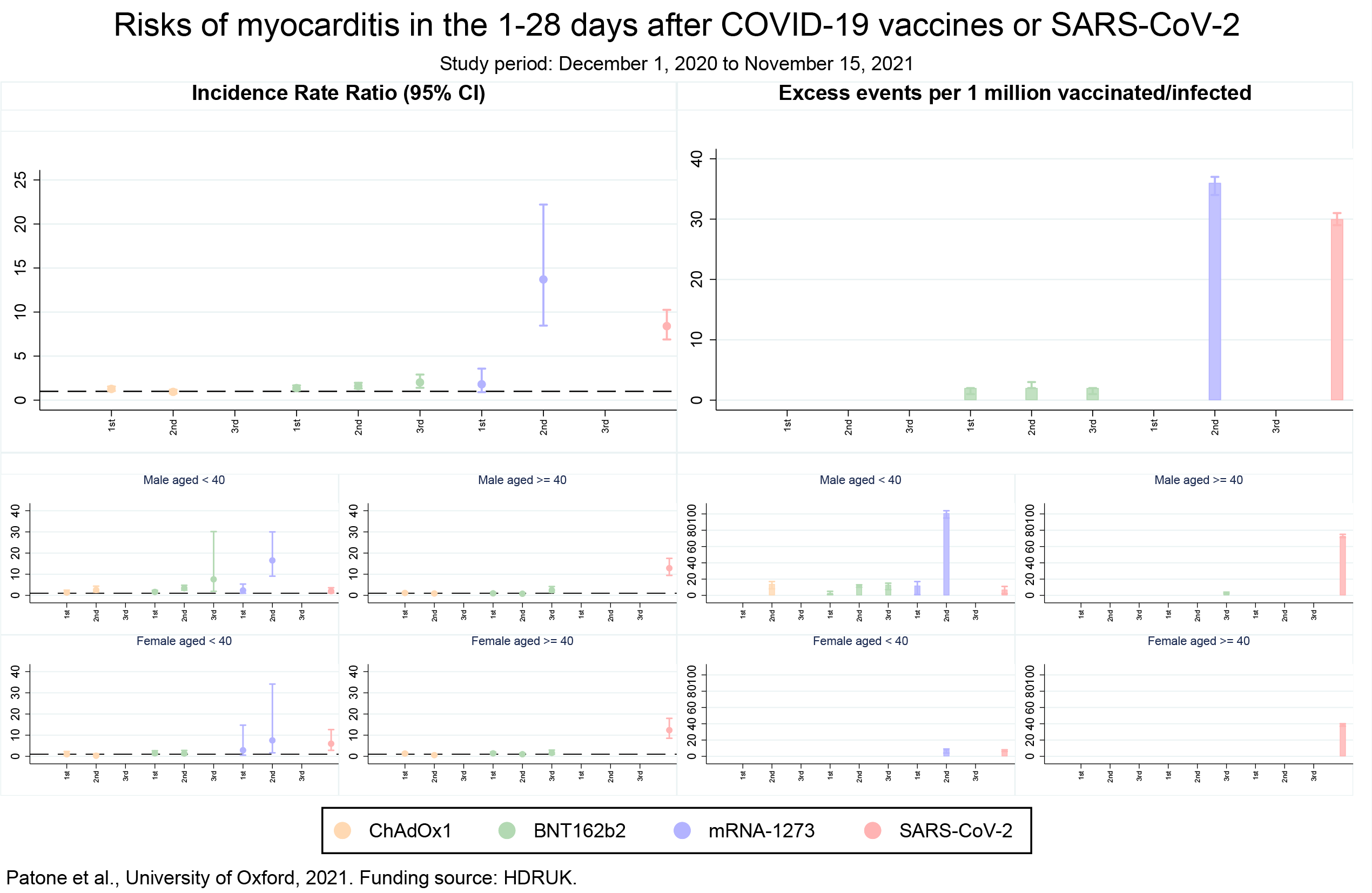
(Left panel) Incidence rate ratios (IRRs) with 95% confidence intervals (CI) and (Right panel) number of excess myocarditis events for million people with 95% confidence intervals (CI) in the 1-28 day risk periods after the first, second and third dose of ChAdOx1, BNT162b2 and mRNA-1273 vaccine or a positive SARS-CoV-2 test in (top) a population of 42,200,614 vaccinated individuals and (bottom) younger males (n=5,893,724), older males (n=11,694,015), younger females (n=6,905,830) and older females (n=13,708,352).

We estimated the number of excess myocarditis events per million persons in the 1-28 days following each exposure for the main analysis and by age and sex (Supplemental Table 5 and Figure 1). Following the first dose of the ChAdOx1 and BNT162b2 vaccines an additional 1 (95%CI 0, 2) and 2 (95%CI 1, 2) myocarditis events per million persons exposed would be anticipated, respectively. Following the second dose of BNT162b2 and mRNA-1273 an additional 2 (95%CI 2, 3) and 36 (95%CI 34, 37) myocarditis events would be anticipated, respectively. Following a third dose of BNT162b2 an additional 2 (95%CI 1, 2) myocarditis events per million persons would be anticipated. These estimates compare to an additional 30 (95%CI 29, 31) myocarditis events per million in the 1-28 days following a SARS-CoV-2 positive test.

In males aged less than 40 years, we estimated an additional 3 (95%CI 1, 5) and 12 (95%CI 1, 13) myocarditis events per million in the 1-28 days following a first dose of BNT162b2 and mRNA-1273, respectively; an additional 14 (95%CI 8, 17), 12 (95%CI 1, 7) and 101 (95%CI 95, 104) myocarditis events following a second dose of ChAdOx1, BNT162b2 and mRNA-1273, respectively; and an additional 13 (95%CI 7, 15) myocarditis events following a third dose of BNT162b2 vaccine. This compares with 7 (95%CI 2, 11) additional myocarditis events in the 1-28 days following a positive SARS-CoV-2 test. In older males, we estimated 3 (95% CI 2, 4) and 73 (95% 71, 75) additional myocarditis events per million following a third dose of BNT162b2 and a positive SARS-CoV-2 test, respectively.

In females aged less than 40 years, we estimated an additional 8 (95% CI 4, 9) and 7 (95% CI 6, 8) events per million following a second dose of mRNA-1273 and a positive SARS-CoV-2 test, respectively. In older females, we estimated no additional myocarditis events following vaccination, but an additional 39 (95% CI 37, 40) events per million following a positive SARS-CoV-2 test.

We report several observations that may have implications for policy makers and the public. First, we confirm and extend our previous findings in more than 42 million persons that the risk of hospitalization or death from myocarditis following COVID-19 infection is higher than the risk associated with vaccination in the overall population. Second, the risk of myocarditis is greater following sequential doses of mRNA vaccine than sequential doses of the adenovirus vaccine. For the first time, we observe an increase in myocarditis events following a third dose of BNT162b vaccine. Whilst the incidence rate ratios are higher sequentially following each dose of mRNA vaccine, the risk remains small in the overall population with an estimated 2 additional cases of myocarditis per million following a booster dose of BNT162b. Third, we report the risk associated with vaccination and infection in younger persons stratified by sex. Despite more myocarditis events occurring in older persons, the risk following COVID-19 vaccination was largely restricted to younger males aged less than 40 years, where the risks of myocarditis following vaccination and infection were similar. However, the notable exception was that in younger males receiving a second dose of mRNA-1273 vaccine, the risk of myocarditis was higher following vaccination than infection, with an additional 101 events estimated following a second dose of mRNA-1273 vaccine compared to 7 events following a positive SARS-CoV-2 test.

There are some limitations we should acknowledge. First, the number of people receiving a third dose of ChAdOx1, or mRNA-1273 vaccine was too small to evaluate the risk of myocarditis. Second, we relied on hospital admission codes and death certification to define myocarditis, and it is possible that we have over or underestimated risk, due to misclassification. Third, although we were able to include 2,136,189 children aged 13 to 17 years old in this analysis, the number of myocarditis events was too small (n=43 in all periods and n=15 in the 1-28 days post vaccination) in this population and precluded an evaluate of risk. Given our observation that risk is largely confined to males under the age of 40 years further research is needed pooling data from international studies to evaluate further the risks in children.

In summary, the risk of hospital admission or death from myocarditis is greater following COVID-19 infection than following vaccination and remains modest following sequential doses of mRNA vaccine including a third booster dose of BNT162b in the overall population. However, the risk of myocarditis following vaccination is consistently higher in younger males, particularly following a second dose of RNA mRNA-1273 vaccine.

## Data Availability

The data that support the findings of this study - National Immunisation (NIMS) Database of COVID-19, mortality (ONS), hospital admissions (HES) and SARS-CoV-2 infection data (PHE) -are not publicly available because they are based on de-identified national clinical records. Due to national and organizational data privacy regulations, individual-level data such as those used for this study cannot be shared openly.

## ACKNOWLEDGEMENTS

This project involves data derived from patient-level information collected by the NHS, as part of the care and support of cancer patients. The SARS-Cov-2 test data are collated, maintained and quality assured by Public Health England (PHE). Access to the data was facilitated by the PHE Office for Data Release. The Hospital Episode Statistics, Secondary Users Service (SUS-PLUS) datasets and civil registration data are used by permission from NHS Digital who retain the copyright in that data. NHS Digital and Public Health England bears no responsibility for the analysis or interpretation of the data. KK is supported by the National Institute for Health Research (NIHR) Applied Research Collaboration East Midlands (ARC-EM) and NIHR Lifestyle BRC. MS-H is supported by the National Institute for Health Research Clinician Scientist Award (NIHR-CS-2016-16-011). JHC and KMC are supported by the NIHR Oxford Biomedical Research Centre. NLM and KMC are supported by the British Heart Foundation (Chair Awards CH/F/21/90010, CH/16/1/32013), Programme Grant (RG/20/10/34966) and Research Excellence Awards (RE/18/5/34216, RE18/3/34214). AS is supported by the Health Data Research UK BREATHE Hub. This research is part of the Data and Connectivity National Core Study, led by Health Data Research UK in partnership with the office of National Statistics and funded by UK Research and Innovation (grant ref MC_PC_20029). The investigators acknowledge the philanthropic support of the donors to the University of Oxford’s COVID-19 Research Response Fund. The views expressed in this publication are those of the author(s) and not necessarily those of the NHS, the UK NIHR or the Department of Health. The funders of this study had no role in the design and conduct of the study and did not review or approve the manuscript. The views expressed are those of the authors and not necessarily the funders. MP, JHC and CC had full access to all the study data and JHC had final responsibility for submission. This project is supported by a patient and public involvement advisory panel which we thank for its continued support and guidance. The input of the panel has helped us identify priority questions for consideration and also supported analysis. PPIE advisers were supportive of the vital importance of reporting on cardiac risks associated with both vaccination against COVID-19 and COVID-19 itself.

## AUTHOR CONTRIBUTIONS

MP, JHC, CC led the study conceptualisation, development of the research question and analysis plan. JHC obtained funding, designed the analysis, obtained data approvals, contributed to interpretation of the analysis. MP undertook the data specification, curation, analysis. MP and NLM wrote the first draft of the paper. SD undertook and reported on the PPIE engagement. LH, KMC, FZ, XWM, NLM, KK, MSH, PW, AH, FZ, SD, AS contributed to the discussion on protocol development and provided critical feedback on drafts of the manuscript. All authors approved the protocol, contributed to the critical revision of the manuscript, and approved the final version of the manuscript.

## DECLARATION OF INTERESTS

AS is a member of the Scottish Government Chief Medical Officer’s COVID-19 Advisory Group, the Scottish Government’s Standing Committee on Pandemics. and AstraZeneca’s Thrombotic Thrombocytopenic Advisory Group. All roles are unremunerated.

JHC reports grants from National Institute for Health Research (NIHR) Biomedical Research Centre, Oxford, grants from John Fell Oxford University Press Research Fund, grants from Cancer Research UK (CR-UK) grant number C5255/A18085, through the Cancer Research UK Oxford Centre, grants from the Oxford Wellcome Institutional Strategic Support Fund (204826/Z/16/Z) and other research councils, during the conduct of the study. JHC is an unpaid director of QResearch, a not-for-profit organisation which is a partnership between the University of Oxford and EMIS Health who supply the QResearch database used for this work. JHC is a founder and shareholder of ClinRisk ltd and was its medical director until 31^st^ May 2019. ClinRisk Ltd produces open and closed source software to implement clinical risk algorithms (outside this work) into clinical computer systems. JHC is chair of the NERVTAG risk stratification subgroup and a member of SAGE COVID-19 groups and the NHS group advising on prioritisation of use of monoclonal antibodies in SARS-CoV-2 infection.

AH is a member of the Joint Committee on Vaccination and Immunisation (JCVI).

KK is a member of the Governments Scientific Advisory Group for Emergencies.

All other authors declare no competing interests related to this paper.

## ONLINE METHODS

### Data

We used the National Immunisation (NIMS) Database of COVID-19 vaccination to identify vaccine exposure. This includes vaccine type, date and doses for all people vaccinated in England. We linked NIMS vaccination data, at individual level, to national data for mortality (ONS), hospital admissions (HES) and SARS-CoV-2 infection data (SGSS).

### Study design

The self-controlled case series (SCCS) design was used, this design was originally developed to examine vaccine safety [13, 14]. The analyses are conditional on each case, so any fixed characteristics during the study period, such as sex, age, ethnicity or chronic conditions, are inherently controlled for. Any time-varying factors, like seasonal variation, need to be adjusted for in the analyses.

### Study period and population

People were considered eligible for inclusion if they were at least 13 years old and had received at least one dose of ChAdOx1 (AstraZeneca), BNT162b2 (Pfizer) and mRNA-1273 (Moderna) and were admitted to hospital with or died from myocarditis between December 1 2020 and November 15 2021 (last data update). Patients were followed up from the study start (December 1 2020) to the earliest of the end of the study period (November 15 2021) or when they died. People were excluded if they had a hospital admission for myocarditis in the two years prior to the start of the study period or if they received Ad26.COV2.S (Janssen) vaccine as there were too few doses delivered to permit a meaningful analysis.

### Outcomes

The outcomes of interest in this study were hospital admission or death from myocarditis. Myocarditis was defined as the first hospital admission in the study period or death using International Classification of Diseases (ICD)-10 codes (Supplementary Table 6).

### Exposures

The exposure variable included the first, second and third dose of the ChAdOx1, BNT162b2 and mRNA-1273 vaccines. Infection with SARS-CoV-2 defined as a COVID-19 reverse transcription– polymerase chain reaction (RT-PCR) positive test was included as a separated exposure variable. Only the first positive test within the study period was used. We defined the exposure risk intervals as the following pre-specified time-periods: 0, 1-7, 8-14, 15-21 and 22-28 days after each exposure date, under the assumption that the adverse events under consideration are unlikely to be related to exposure from 28 days post-exposure. People who experience the outcome are likely to delay vaccination until symptoms have improved, and therefore a pre-risk period of 1-28 days before each exposure was removed from the baseline period to account for this potential bias. Hospital admission for myocarditis often results in testing for SARS-CoV-2. Whilst these outcomes may well be caused by SARS-CoV-2 infection, reverse causality involved in their detection could over- or under-estimate the effect of infection on myocarditis. To interrogate this potential source of bias, we allocated day 0 to a risk period of its own.

### Statistical analysis

We described characteristics of the whole vaccinated cohort in terms of age, sex, ethnicity, SARS-CoV-2 positive test status, number of doses received, homologous and heterologous vaccination by vaccine doses and type.

We described demographic characteristics of vaccinated people with admission or death from myocarditis during and outwith the risk period (1-28 days post each vaccine dose).

The SCCS models were fitted using a conditional Poisson regression model. Incidence rate ratios (IRR), the relative rate ratio of hospital admissions or deaths due to each outcome of interest in risk periods relative to baseline periods, were estimated by the SCCS model adjusted for two-week calendar periods as time-varying covariates (to account for seasonal effects).

Absolute risk differences cannot be obtained using SCCS. We supplemented our estimates of IRRs with measures of effect of each exposure in absolute terms using a method developed to estimate the number of exposures needed to produce one excess adverse outcome and the excess number of events per 1,000,000 exposed for each outcome [15].

Stata version 17 was used for these analyses.

### Code availability

The code used for this study has been deposited in the git repository of the research group, which is protected by privacy. Access to the code is available from the authors on request for non-commercial, academic and research use only. Stata version 17 was used for these analyses.

## Notes

### Clinical Protocols

https://www.qresearch.org/media/1304/ox107_covid_vaccine_safety_protocol.pdf

### Author Declarations

National Health Service Research Ethics Committee (NHS REC) approval was obtained from East Midlands-Derby Research Ethics Committee [reference 04/03/2021].

