## Supplementary tables for "Risk of myocarditis following sequential COVID-19 vaccinations by age and sex"

**Supplementary Table 1. Baseline demographic characteristics of people receiving either ChAdOx1, BNT162b2 or mRNA-1273 vaccines or testing positive to SARS-CoV-2 virus (in those vaccinated with either vaccine), in England between 1 December 2020 until 15 November 2021. Figures are column % (counts).**

| | ChAdOx1 | BNT162b2 | mRNA-1273 | ChAdOx1 | BNT162b2 | mRNA-1273 | ChAdOx1 | BNT162b2 | mRNA-1273 | Positive SARS-CoV-2 test* |
|---|---|---|---|---|---|---|---|---|---|---|
| | 1 dose (n = 42,200,614) | | | 2 dose (n = 38,347,981) | | | 3 dose (n = 10,978,507) | | | (n = 5,185,772) |
| | *Col % (counts)* | *Col % (counts)* | *Col % (counts)* | *Col % (counts)* | *Col % (counts)* | *Col % (counts)* | *Col % (counts)* | *Col % (counts)* | *Col % (counts)* | *Col % (counts)* |
| Total number of people | 20,646,456 | 20,391,600 | 1,162,558 | 20,059,058 | 17,294,004 | 1,039,919 | 35,608 | 10,599,183 | 343,716 | 5,185,772 |
| **Sex** | | | | | | | | | | |
| Women | 49.3 (10182140) | 49.0 (9985535) | 38.4 (446579) | 49.4 (9905031) | 50.1 (8657542) | 39.0 (405748) | 57.2 (20369) | 56.5 (5987783) | 51.6 (177238) | 52.2 (2707945) |
| Men | 43.1 (8900093) | 40.2 (8199395) | 42.0 (488323) | 43.2 (8657222) | 39.6 (6849358) | 41.8 (434478) | 37.4 (13331) | 40.3 (4274761) | 44.2 (151816) | 40.4 (2095960) |
| Not recorded | 7.6 (1564224) | 10.8 (2206669) | 19.6 (227656) | 7.5 (1496805) | 10.3 (1787103) | 19.2 (199693) | 5.4 (1908) | 3.2 (336639) | 4.3 (14662) | 7.4 (381867) |
| **Age** | | | | | | | | | | |
| Mean age (SD) | 54.9 (14.8) | 43.6 (22.3) | 32.1 (9.4) | 55.0 (14.7) | 47.3 (21.6) | 32.4 (9.4) | 62.5 (17.2) | 66.7 (15.4) | 63.2 (12.4) | 42.0 (18.2) |
| 13-17 years | 0.0 (10212) | 9.3 (1887751) | 0.0 (556) | 0.0 (9064) | 1.3 (220744) | 0.0 (248) | 0.1 (22) | 0.1 (7422) | 0.0 (170) | 7.1 (367113) |
| 18-29 years | 5.2 (1080573) | 24.6 (5009612) | 43.2 (502147) | 5.1 (1020629) | 24.8 (4281647) | 41.5 (431451) | 4.2 (1495) | 2.8 (296943) | 2.2 (7516) | 22.2 (1151541) |
| 30-39 years | 7.9 (1633888) | 21.8 (4449421) | 36.0 (418848) | 7.7 (1553370) | 23.3 (4034166) | 36.7 (381825) | 6.3 (2250) | 4.3 (455032) | 3.6 (12351) | 18.0 (931836) |
| 40-49 years | 22.1 (4563009) | 8.6 (1752175) | 18.6 (216106) | 22.0 (4404792) | 9.3 (1615419) | 19.7 (204584) | 11.9 (4229) | 6.5 (692898) | 6.3 (21712) | 19.0 (983809) |

| | | | | | | | | | | |
|---|---|---|---|---|---|---|---|---|---|---|
| 50-59 years | 27.5 (5673107) | 8.2 (1665317) | 1.5 (17748) | 27.6 (5543761) | 9.3 (1608620) | 1.4 (14750) | 19.2 (6845) | 13.4 (1424029) | 17.3 (59467) | 16.9 (878706) |
| 60-69 years | 19.8 (4083537) | 8.7 (1766207) | 0.4 (5184) | 20.0 (4012352) | 10.0 (1737947) | 0.5 (4847) | 18.2 (6498) | 21.0 (2223266) | 39.6 (136187) | 8.9 (462660) |
| 70-79 years | 13.4 (2762915) | 9.7 (1973783) | 0.1 (1542) | 13.5 (2717662) | 11.3 (1952441) | 0.2 (1700) | 24.1 (8586) | 32.9 (3492004) | 26.0 (89442) | 4.5 (235613) |
| 80-89 years | 3.1 (630417) | 7.9 (1619014) | 0.0 (369) | 3.0 (605236) | 9.2 (1587386) | 0.0 (438) | 11.9 (4239) | 16.2 (1718395) | 4.1 (14157) | 2.5 (127574) |
| 90+ years | 1.0 (208754) | 1.3 (268204) | 0.0 (56) | 1.0 (192180) | 1.5 (255591) | 0.0 (75) | 4.1 (1444) | 2.7 (289173) | 0.8 (2714) | 0.9 (46910) |
| Not recorded | 0.0 (301) | 5.1 (1030662) | 0.0 (47) | 0.0 (107) | 0.1 (13228) | 0.0 (11) | < 0.1 (1) | 0.0 (102) | | 4.0 (208182) |
| **Ethnicity** | | | | | | | | | | |
| White | 67.8 (13998166) | 63.7 (12989545) | 53.3 (619541) | 68.0 (13632603) | 64.4 (11139124) | 53.7 (558043) | 73.4 (26148) | 76.7 (8131980) | 76.2 (261992) | 66.8 (3461571) |
| Other ethnic groups | 8.1 (1672963) | 9.5 (1936026) | 7.6 (88043) | 8.0 (1601621) | 8.7 (1508302) | 7.2 (75218) | 7.9 (2808) | 5.8 (618083) | 4.0 (13747) | 10.6 (549690) |
| Ethnicity not recorded | 24.1 (4975328) | 26.8 (5466028) | 39.1 (454974) | 24.1 (4824834) | 26.9 (4646577) | 39.1 (406658) | 18.7 (6652) | 17.4 (1849120) | 19.8 (67977) | 22.6 (1174511) |
| **History of myocarditis** | | | | | | | | | | |
| Prior myocarditis | 0.0 (1836) | 0.0 (1608) | 0.0 (65) | 0.0 (1776) | 0.0 (1482) | 0.0 (56) | 0.0 (10) | 0.0 (1289) | 0.0 (38) | 0.0 (623) |
| **COVID-19 status** | | | | | | | | | | |
| No covid | 87.9 (18150536) | 87.5 (17849752) | 87.3 (1014636) | 88.0 (17648006) | 88.7 (15341130) | 87.8 (913362) | 88.6 (31559) | 92.6 (9812390) | 93.0 (319628) | - |
| Covid prior vaccination | 5.9 (1226752) | 7.4 (1513590) | 8.1 (94199) | 5.9 (1181865) | 6.3 (1083950) | 7.7 (79838) | 6.5 (2329) | 4.0 (422009) | 3.5 (12122) | 54.7 (2834579) |
| Covid after 1st dose | 0.7 (136878) | 2.6 (525639) | 3.1 (36443) | 0.5 (97652) | 2.1 (365674) | 2.8 (29318) | 0.8 (296) | 0.7 (75604) | 0.3 (1088) | 13.5 (698993) |

| | | | | | | | | | | |
|---|---|---|---|---|---|---|---|---|---|---|
| Covid after 2nd dose | 5.4 (1111972) | 2.3 (474831) | 1.5 (17274) | 5.5 (1111209) | 2.7 (475472) | 1.7 (17393) | 1.1 (390) | 2.3 (242891) | 2.9 (10092) | 30.9 (1604087) |
| Covid after 3rd dose | 0.1 (20319) | 0.1 (27787) | < 0.1 (6) | 0.1 (20326) | 0.2 (27777) | < 0.1 (8) | 2.9 (1034) | 0.4 (46289) | 0.2 (786) | 0.9 (48113) |
| **Number of doses** | | | | | | | | | | |
| One dose | 2.6 (538297) | 15.4 (3145304) | 10.7 (124782) | - | - | - | - | - | - | 13.4 (695253) |
| Two doses | 73.4 (15145207) | 55.1 (11231447) | 89.2 (1037051) | 75.2 (15091876) | 65.3 (11284348) | 99.9 (1038375) | - | - | - | 70.9 (3675578) |
| Three doses | 24.0 (4962953) | 29.5 (6014848) | 0.1 (725) | 24.8 (4967182) | 34.7 (6009655) | 0.1 (1544) | 100.0 (35608) | 100.0 (10599183) | 100.0 (343716) | 15.7 (814941) |
| **Type of vaccines** | | | | | | | | | | |
| Two doses of ChAdOx1 | 97.0 (20018696) | - | - | 99.8 (20018696) | - | - | 82.1 (29244) | 44.3 (4692400) | 66.7 (229396) | 46.4 (2406251) |
| Two doses of BNT162b2 | - | 84.4 (17203272) | | - | 99.5 (17203272) | - | 5.6 (1994) | 55.5 (5882812) | 33.0 (113477) | 37.4 (1940244) |
| Two doses of mRNA-1273 | - | - | 88.9 (1033088) | - | - | 99.3 (1033088) | < 0.1 (5) | 0.0 (320) | 0.1 (259) | 2.4 (125757) |

*amongst vaccinated

**Supplementary Table 2: Demographic and clinical characteristics of patients who were admitted to hospital for myocarditis in the 28 days following a COVID-19 vaccine first, second dose and third dose or SARS-CoV-2 infection amongst the vaccinated population in England from 1 December 2020 until 15 November 2021 (cells with < 5 are suppressed).**

| | **Myocarditis** | | | | | | | | | | |
|---|---|---|---|---|---|---|---|---|---|---|---|
| | **Baseline** | **Risk set (1-28 days post)** | | | | | | | | | |
| | | **ChAdOx1 (1st dose)** | **ChAdOx1 (2nd dose)** | **ChAdOx1 (3rd dose)** | **BNT162b2 (1st dose)** | **BNT162b2 (2nd dose)** | **BNT162b2 (3rd dose)** | **mRNA-1273 (1st dose)** | **mRNA-1273 (2nd dose)** | **mRNA-1273 (3rd dose)** | **Positive SARS-CoV-2 test** |
| Total number of people | 1696 | 139 | 89 | 0 | 120 | 114 | 39 | 11 | 40 | 0 | 177 |
| **Sex** | | | | | | | | | | | |
| Women | 41.7 (697) | 41.7 (58) | 27.0 (24) | - | 42.5 (51) | 29.8 (34) | 41.0 (16) | * | * | - | 40.7 (72) |
| Men | 58.7 (995) | 58.3 (81) | 73.0 (65) | - | 56.7 (68) | 70.2 (80) | 59.0 (23) | 81.8 (9) | 90.0 (36) | - | 59.3 (105) |
| **Age** | | | | | | | | | | | |
| Mean age (SD) | 53.8 (19.6) | 57.5 (17.6) | 54.2 (18.1) | - | 49.7 (24.1) | 45.9 (25.0) | 68.4 (17.4) | 27.0 (9.5) | 24.9 (6.3) | - | 61.5 (17.9) |
| <40 years | 26.2 (444) | 14.4 (20) | 25.8 (23) | - | 44.2 (53) | 57.0 (65) | * | 90.9 (10) | 97.5 (39) | - | 14.1 (25) |
| >=40 years | 73.8 (1252) | 85.6 (119) | 74.2 (66) | - | 55.8 (67) | 43.0 (49) | 92.3 (36) | * | * | - | 85.9 (152) |
| **Deaths** | 9.1 (155) | 24.5 (34) | 7.9 (7) | - | 15.0 (18) | 10.5 (12) | 20.5 (8) | - | - | - | 6.8 (12) |
| **Covid Status** | | | | | | | | | | | |
| No covid | - | 74.1 (103) | 84.3 (75) | - | 73.3 (88) | 91.2 (104) | 84.6 (33) | 54.5 (6) | 90.0 (36) | - | - |
| Covid prior vaccination | - | 12.9 (18) | 11.2 (10) | - | 10.0 (12) | * | * | * | * | - | 63.8 (113) |
| Covid after 1st dose | - | 10.8 (15) | * | - | 14.2 (17) | * | - | * | * | - | 15.8 (28) |

| | | | | | | | | | | | |
|---|---|---|---|---|---|---|---|---|---|---|---|
| Covid after 2nd dose | - | * | * | - | * | * | * | - | - | - | 20.3 (36) |
| Covid after 3rd dose | - | - | - | - | * | - | - | - | - | - | - |
| One dose | - | 44.6 (62) | - | - | 53.3 (64) | - | - | 90.9 (10) | - | - | 15.3 (27) |
| Two doses | - | 41.0 (57) | 78.7 (70) | - | 25.0 (30) | 76.3 (87) | - | * | 100.0 (40) | - | 63.8 (113) |
| Three doses | - | 14.4 (20) | 21.3 (19) | - | 21.7 (26) | 23.7 (27) | 100.0 (39) | - | - | - | 20.9 (37) |
| Two doses of ChAdOx1 | - | 51.8 (72) | 98.9 (88) | - | - | - | 38.5 (15) | - | - | - | 53.1 (94) |
| Two doses of BNT162b2 | - | - | - | - | 44.2 (53) | 99.1 (113) | 61.5 (24) | - | - | - | 29.9 (53) |
| Two doses of mRNA-1273 | - | - | - | - | - | - | - | * | 100.0 (40) | - | * |
| Lag between first and second doses >= 28 days | - | 54.7 (76) | 100.0 (89) | - | 45.8 (55) | 96.5 (110) | 92.3 (36) | * | 100.0 (40) | - | 84.2 (149) |

**Supplementary Table 3: Incidence rate ratios (IRR 95% CI) for the main analysis and by age group (aged 40 or younger, older than 40) and sex (female and male) for the outcomes in pre-defined risk periods immediately before and after exposure to vaccination and before and after a positive SARS-CoV-2 test result, adjusted for calendar time from December 1 2020 to November 15 2021 (cells with * are suppressed).**

| | **ChAdOx1** | **BNT162b2** | **mRNA-1273** | **ChAdOx1** | **BNT162b2** | **mRNA-1273** | **ChAdOx1** | **BNT162b2** | **mRNA-1273** | **Positive SARS-CoV-2 test*** |
|---|---|---|---|---|---|---|---|---|---|---|
| | **1 dose (n = 42,200,614)** | | | **2 dose (n = 38,347,981)** | | | **3 dose (n = 10,978,507)** | | | **(n = 5,185,772)** |
| | *Col % (counts)* | *Col % (counts)* | *Col % (counts)* | *Col % (counts)* | *Col % (counts)* | *Col % (counts)* | *Col % (counts)* | *Col % (counts)* | *Col % (counts)* | *Col % (counts)* |
| Total number of people | 20,646,456 | 20,391,600 | 1,162,558 | 20,059,058 | 17,294,004 | 1,039,919 | 35,608 | 10,599,183 | 343,716 | 5,185,772 |

| | | | | | | | | | | |
|---|---|---|---|---|---|---|---|---|---|---|
| **Sex & Age subgroup** | | | | | | | | | | |
| Male aged < 40 years old | 992,510 | 4,517,315 | 383,899 | 943,070 | 3,282,074 | 336,105 | 1,236 | 199,914 | 6,165 | 949,941 |
| Male aged >= 40 years old | 7,907,552 | 3,682,041 | 104,422 | 7,714,148 | 3,567,272 | 98,372 | 12,095 | 4,074,840 | 135,651 | 1,146,015 |
| Female aged < 40 years old | 1,502,095 | 5,054,889 | 348,846 | 1,427,406 | 3,847,428 | 312,891 | 2,141 | 502,825 | 12,763 | 1,256,488 |
| Female aged >= 40 years old | 8,680,029 | 4,930,590 | 97,733 | 8,477,618 | 4,810,093 | 92,857 | 18,228 | 5,484,946 | 164,475 | 1,451,451 |

**Supplementary Table 4: Incidence rate ratios (IRR 95% CI) for the main analysis and by age group (aged 40 or younger, older than 40) and sex (female and male) for the outcomes in pre-defined risk periods immediately before and after exposure to vaccination and before and after a positive SARS-CoV-2 test result, adjusted for calendar time from 1 December 2020 to 15 November 2021 (cells with * are suppressed).**

| | **ChAdOx1nCoV-19 vaccine** | | **BNT162b2 mRNA vaccine** | | **mRNA-1273 vaccine** | | **Positive SARS-CoV-2 test** | |
|---|---|---|---|---|---|---|---|---|
| **Time period** | **events** | | **IRR (95% CI)** | | **events** | | **IRR (95% CI)** | |
| **Main analysis** | | | | | | | | |
| Baseline* | 1696 | 1.00 | 1696 | 1.00 | 1696 | 1.00 | 2268 | 1.00 |
| Pre-risk** | 283 | 0.71 (0.62, 0.81) | 283 | 0.71 (0.62, 0.81) | 283 | 0.71 (0.62, 0.81) | 43 | 2.20 (1.58, 3.06) |
| 0 day: 1st dose/positive test | * | 0.55 (0.14, 2.20) | * | 1.05 (0.34, 3.27) | * | n/a | 51 | 68.70 (50.50, 93.47) |
| 1-7 days: 1st dose/positive test | **47** | **1.77 (1.30, 2.41)** | **35** | **1.64 (1.16, 2.32)** | **8** | **6.09 (2.82, 13.11)** | 89 | 17.28 (13.46, 22.18) |
| 8-14 days: 1st dose/positive test | 34 | 1.21 (0.85, 1.72) | 31 | 1.34 (0.93, 1.94) | * | 1.03 (0.23, 4.57) | 47 | 9.20 (6.70, 12.63) |
| 15-21 days: 1st dose/positive test | 29 | 1.02 (0.70, 1.50) | 25 | 1.08 (0.72, 1.62) | * | n/a | 23 | 4.53 (2.94, 6.97) |
| 22-28 days: 1st dose/positive test | 29 | 1.03 (0.70, 1.50) | 29 | 1.33 (0.91, 1.94) | * | n/a | 18 | 3.36 (2.07, 5.44) |
| 0 day: 2nd dose | * | 0.60 (0.15, 2.40) | * | n/a | * | n/a | | |
| 1-7 days: 2nd dose | 14 | 0.60 (0.35, 1.02) | **48** | **2.64 (1.96, 3.56)** | **36** | **46.73 (28.58, 76.42)** | | |
| 8-14 days: 2nd dose | 29 | 1.23 (0.84, 1.79) | **27** | **1.51 (1.03, 2.23)** | * | n/a | | |
| 15-21 days: 2nd dose | 19 | 0.80 (0.51, 1.27) | 24 | 1.35 (0.90, 2.04) | * | 2.79 (0.66, 11.70) | | |
| 22-28 days: 2nd dose | 27 | 1.13 (0.76, 1.66) | 15 | 0.84 (0.50, 1.41) | * | n/a | | |

| | | | | | | | | |
|---|---|---|---|---|---|---|---|---|
| 0 day: 3rd dose | * | n/a | * | n/a | * | n/a | | |
| 1-7 days: 3rd dose | * | n/a | 12 | 1.64 (0.91, 2.96) | * | n/a | | |
| 8-14 days: 3rd dose | * | n/a | **14** | **2.48 (1.42, 4.33)** | * | n/a | | |
| 15-21 days: 3rd dose | * | n/a | **12** | **3.02 (1.65, 5.52)** | * | n/a | | |
| 22-28 days: 3rd dose | * | n/a | * | n/a | * | n/a | | |
| **Age <40 & Male** | | | | | | | | |
| Baseline* | 304 | 1.00 | 304 | 1.00 | 304 | 1.00 | 484 | 1.00 |
| Pre-risk** | 34 | 0.61 (0.42, 0.88) | 34 | 0.61 (0.42, 0.88) | 34 | 0.61 (0.42, 0.88) | 9 | 1.33 (0.65, 2.69) |
| 0 day: 1st dose/positive test | * | n/a | * | n/a | * | n/a | **8** | **29.82 (14.06, 63.26)** |
| 1-7 days: 1st dose/positive test | **8** | **3.26 (1.53, 6.91)** | **16** | **2.98 (1.75, 5.07)** | **6** | **7.97 (3.17, 20.05)** | **8** | **4.63 (2.19, 9.78)** |
| 8-14 days: 1st dose/positive test | * | 0.80 (0.19, 3.28) | **13** | **2.20 (1.22, 3.94)** | * | 2.05 (0.46, 9.16) | * | n/a |
| 15-21 days: 1st dose/positive test | * | 1.24 (0.38, 3.99) | * | 0.35 (0.09, 1.41) | * | n/a | * | n/a |
| 22-28 days: 1st dose/positive test | * | n/a | 8 | 1.42 (0.69, 2.93) | * | n/a | * | 2.25 (0.81, 6.22) |
| 0 day: 2nd dose | * | n/a | * | n/a | * | n/a | | |
| 1-7 days: 2nd dose | * | 1.14 (0.27, 4.73) | **33** | **8.05 (5.37, 12.06)** | **32** | **54.65 (29.74, 100.40)** | | |
| 8-14 days: 2nd dose | **10** | **5.36 (2.67, 10.78)** | **9** | **2.24 (1.13, 4.46)** | * | n/a | | |
| 15-21 days: 2nd dose | * | 1.01 (0.24, 4.17) | **10** | **2.46 (1.27, 4.74)** | * | 3.57 (0.82, 15.60) | | |
| 22-28 days: 2nd dose | **7** | **3.21 (1.43, 7.22)** | * | 0.94 (0.35, 2.57) | * | n/a | | |
| 0 day: 3rd dose | * | n/a | * | n/a | * | n/a | | |
| 1-7 days: 3rd dose | * | n/a | * | n/a | * | n/a | | |
| 8-14 days: 3rd dose | * | n/a | * | n/a | * | n/a | | |
| 15-21 days: 3rd dose | * | n/a | * | n/a | * | n/a | | |
| 22-28 days: 3rd dose | * | n/a | * | n/a | * | n/a | | |
| **Age >= 40 & Male** | | | | | | | | |
| Baseline* | 691 | 1.00 | 691 | 1.00 | 691 | 1.00 | 897 | 1.00 |
| Pre-risk** | 148 | 0.81 (0.67, 0.99) | 148 | 0.81 (0.67, 0.99) | 148 | 0.81 (0.67, 0.99) | **16** | **2.64 (1.53, 4.57)** |

| | | | | | | | | |
|---|---|---|---|---|---|---|---|---|
| 0 day: 1st dose/positive test | * | n/a | * | n/a | * | n/a | **23** | **100.01 (62.18, 160.83)** |
| 1-7 days: 1st dose/positive test | 20 | 1.43 (0.89, 2.29) | 6 | 0.84 (0.37, 1.91) | * | n/a | **45** | **27.54 (18.87, 40.21)** |
| 8-14 days: 1st dose/positive test | 21 | 1.38 (0.87, 2.21) | * | 0.55 (0.20, 1.50) | * | n/a | **23** | **13.44 (8.38, 21.55)** |
| 15-21 days: 1st dose/positive test | 13 | 0.84 (0.47, 1.49) | 9 | 1.16 (0.58, 2.29) | * | n/a | **13** | **7.37 (4.08, 13.31)** |
| 22-28 days: 1st dose/positive test | 14 | 0.88 (0.51, 1.52) | 10 | 1.24 (0.65, 2.39) | * | n/a | **10** | **5.38 (2.78, 10.42)** |
| 0 day: 2nd dose | * | n/a | * | n/a | * | n/a | | |
| 1-7 days: 2nd dose | 10 | 0.78 (0.41, 1.49) | 5 | 0.65 (0.27, 1.59) | * | n/a | | |
| 8-14 days: 2nd dose | 13 | 1.05 (0.59, 1.84) | 5 | 0.65 (0.27, 1.60) | * | n/a | | |
| 15-21 days: 2nd dose | 11 | 0.92 (0.50, 1.69) | 7 | 0.93 (0.44, 2.00) | * | n/a | | |
| 22-28 days: 2nd dose | 10 | 0.87 (0.46, 1.64) | 7 | 0.92 (0.43, 1.97) | * | n/a | | |
| 0 day: 3rd dose | * | n/a | * | n/a | * | n/a | | |
| 1-7 days: 3rd dose | * | n/a | * | 0.99 (0.31, 3.15) | * | n/a | | |
| 8-14 days: 3rd dose | * | n/a | **10** | **4.41 (2.22, 8.73)** | * | n/a | | |
| 15-21 days: 3rd dose | * | n/a | **6** | **3.83 (1.61, 9.08)** | * | n/a | | |
| 22-28 days: 3rd dose | * | n/a | * | n/a | * | n/a | | |
| **Age <40** & **Female** | | | | | | | | |
| Baseline* | 140 | 1.00 | 140 | 1.00 | 140 | 1.00 | 176 | 1.00 |
| Pre-risk** | 13 | 0.43 (0.24, 0.77) | 13 | 0.43 (0.24, 0.77) | 13 | 0.43 (0.24, 0.77) | * | n/a |
| 0 day: 1st dose/positive test | * | n/a | * | **6.41 (1.55, 26.51)** | * | n/a | * | **37.07 (10.55, 130.19)** |
| 1-7 days: 1st dose/positive test | **5** | **2.86 (1.07, 7.60)** | * | 1.36 (0.42, 4.39) | * | n/a | **5** | **10.09 (3.67, 27.70)** |
| 8-14 days: 1st dose/positive test | * | n/a | **6** | **2.51 (1.05, 5.98)** | * | n/a | * | n/a |
| 15-21 days: 1st dose/positive test | * | n/a | * | 1.85 (0.66, 5.17) | * | n/a | * | **8.03 (2.33, 27.69)** |
| 22-28 days: 1st dose/positive test | * | n/a | * | n/a | * | n/a | * | 3.87 (0.88, 16.98) |
| 0 day: 2nd dose | * | n/a | * | n/a | * | n/a | | |
| 1-7 days: 2nd dose | * | n/a | **5** | **3.11 (1.23, 7.86)** | * | **28.49 (6.22, 130.41)** | | |
| 8-14 days: 2nd dose | * | n/a | * | 2.58 (0.92, 7.22) | * | n/a | | |
| 15-21 days: 2nd dose | * | n/a | * | n/a | * | n/a | | |

| | | | | | | | | |
|---|---|---|---|---|---|---|---|---|
| 22-28 days: 2nd dose | * | 1.18 (0.28, 5.00) | * | n/a | * | n/a | | |
| 0 day: 3rd dose | * | n/a | * | n/a | * | n/a | | |
| 1-7 days: 3rd dose | * | n/a | * | n/a | * | n/a | | |
| 8-14 days: 3rd dose | * | n/a | * | n/a | * | n/a | | |
| 15-21 days: 3rd dose | * | n/a | * | n/a | * | n/a | | |
| 22-28 days: 3rd dose | * | n/a | * | n/a | * | n/a | | |
| **Age >= 40 & Female** | | | | | | | | |
| Baseline* | 557 | 1.00 | 557 | 1.00 | 557 | 1.00 | 706 | 1.00 |
| Pre-risk** | 88 | 0.63 (0.49, 0.81) | 88 | 0.63 (0.49, 0.81) | 88 | 0.63 (0.49, 0.81) | **17** | **3.93 (2.25, 6.86)** |
| 0 day: 1st dose/positive test | * | 1.50 (0.37, 6.11) | * | n/a | * | n/a | **17** | **96.00 (55.17, 167.06)** |
| 1-7 days: 1st dose/positive test | 14 | 1.46 (0.83, 2.56) | 10 | 1.40 (0.72, 2.74) | * | n/a | **31** | **25.39 (16.17, 39.87)** |
| 8-14 days: 1st dose/positive test | 11 | 1.09 (0.58, 2.03) | 8 | 1.05 (0.50, 2.20) | * | n/a | **22** | **17.85 (10.79, 29.53)** |
| 15-21 days: 1st dose/positive test | 12 | 1.18 (0.64, 2.15) | 9 | 1.24 (0.62, 2.48) | * | n/a | **6** | **4.87 (2.09, 11.36)** |
| 22-28 days: 1st dose/positive test | 14 | 1.41 (0.80, 2.47) | 11 | 1.73 (0.92, 3.27) | * | n/a | * | 1.63 (0.40, 6.72) |
| 0 day: 2nd dose | * | n/a | * | n/a | * | n/a | | |
| 1-7 days: 2nd dose | * | 0.21 (0.05, 0.86) | 5 | 0.80 (0.33, 1.97) | * | n/a | | |
| 8-14 days: 2nd dose | 6 | 0.63 (0.28, 1.44) | 9 | 1.43 (0.72, 2.82) | * | n/a | | |
| 15-21 days: 2nd dose | 6 | 0.63 (0.28, 1.42) | 7 | 1.11 (0.52, 2.39) | * | n/a | | |
| 22-28 days: 2nd dose | 8 | 0.83 (0.41, 1.70) | * | 0.63 (0.23, 1.70) | * | n/a | | |
| 0 day: 3rd dose | * | n/a | * | n/a | * | n/a | | |
| 1-7 days: 3rd dose | * | n/a | **8** | **2.32 (1.09, 4.94)** | * | n/a | | |
| 8-14 days: 3rd dose | * | n/a | * | 1.09 (0.34, 3.56) | * | n/a | | |
| 15-21 days: 3rd dose | * | n/a | 5 | 2.28 (0.88, 5.93) | * | n/a | | |
| 22-28 days: 3rd dose | * | n/a | * | n/a | * | n/a | | |

*Same for each exposure.

**-28-1 days prior to each vaccine dose or positive test. Same for each exposure.

**Supplementary table 5: Measures of effect of vaccinations and SARS-CoV-2 infections presented in absolute terms, number needed to expose for one excess event; excess events per 1 million exposed. Only significant increased risks were reported over the 1-28 days post exposure. When IRR were not significant over the 1-28 days post-vaccine, absolute measures were not given.**

| | **Excess events per 1,000,000 exposed (95% CI)** | | | | |
|---|---|---|---|---|---|
| | **Main analysis** | Male aged < 40 | Male aged >= 40 | Female aged < 40 | Female aged >= 40 |
| **ChAdOx1** | | | | | |
| 1st dose | 1 (0, 2) | - | - | - | - |
| 2nd dose | - | 14 (8, 17) | - | - | - |
| 3rd dose | - | - | - | - | - |
| **BNT162b2** | | | | | |
| 1st dose | 2 (1, 2) | 3 (1, 5) | - | - | - |
| 2nd dose | 2 (2, 3) | 12 (10, 13) | - | - | - |
| 3rd dose | 2 (1, 2) | 13 (7, 15) | 3 (2, 4) | - | - |
| **mRNA-1273** | | | | | |
| 1st dose | - | 12 (1, 17) | - | - | - |
| 2nd dose | 36 (34, 37) | 101 (95, 104) | - | 8 (4, 9) | - |
| 3rd dose | - | - | - | - | - |
| **SARS-CoV-2** | | | | | |
| Positive test | 30 (29, 31) | 7 (2, 11) | 73 (71, 75) | 7 (6, 8) | 39 (37, 40) |

**Supplementary table 6: ICD-10 codes used to identify cases with each outcome.**

| Outcome | ICD-10 code and description |
|---|---|
| **Myocarditis** | I40 - Acute myocarditis<br>I400 - Infective myocarditis<br>I401 - Isolated myocarditis<br>I408 - Other acute myocarditis<br>I409 - Acute myocarditis, unspecified<br>I41 - Myocarditis in diseases classified elsewhere<br>I410 - Myocarditis in bacterial diseases classified elsewhere<br>I411 - Myocarditis in viral diseases classified elsewhere<br>I412 - Myocarditis in other infectious and parasitic diseases classified elsewhere<br>I418 - Myocarditis in other diseases classified elsewhere<br>I514 - Myocarditis, unspecified |
